# A Feasibility Study of Thermography for Detecting Pressure Injuries Across Diverse Skin Tones

**DOI:** 10.1101/2024.10.14.24315465

**Authors:** Miriam Asare-Baiden, Sharon Eve Sonenblum, Kathleen Jordan, Andrew Chung, Judy Wawira Gichoya, Vicki Stover Hertzberg, Joyce C Ho

## Abstract

Pressure injury (PI) detection is challenging, especially in dark skin tones, due to the unreliability of visual inspection. Thermography may serve as a viable alternative as temperature differences in the skin can indicate impending tissue damage. Although deep learning models hold considerable promise toward reliably detecting PI, existing work fails to evaluate performance on diverse skin tones and varying data collection protocols. We collected a new dataset of 35 participants focused on darker skin tones where temperature differences are induced through cooling and cupping protocols. The dataset includes different cameras, lighting, patient pose, and camera distance. We compare the performance of three convolutional neural network (CNN) models trained on either the thermal or the optical images on all skin tones. Our results suggest thermography-based CNN is robust to data collection protocols. Moreover, the visual explanation often captures the region of interest without requiring explicit bounding box labels.

## Introduction

A pressure injury (PI) refers to “localized damage to the skin and underlying soft tissue, usually occurring over a bony prominence or related to medical devices” [1]. It continues to be a major issue with at least 1 in 10 adults admitted to the hospital developing a PI [2]. Furthermore, PI is linked to a decline in quality of life [3], increased mortality rates [4], extended hospital stays, and a higher likelihood of requiring institutional care after discharge. Identifying warning signs of PI can lead to effective preventive actions and early interventions [5, 6].

Contemporary techniques for assessing PI risk include using the Braden Scale [7, 8] and visual inspection. Visual inspection includes assessing the skin for temperature variations, changes in tissue consistency, and the presence of tenderness. Yet visual inspection for dark skin tone patients is especially challenging and has led to racial disparities in PI outcomes [9]. Given that PI may result in temperature abnormalities from partial or complete capillary occlusion at the site, thermography (i.e., thermal imaging) has been explored as an alternative technological approach for PI detection [10]. Several existing works have suggested deep learning models such as convolutional neural networks (CNNs) can achieve high predictive performance for detecting PI [11, 12, 13]. However, these studies suffer from 3 major limitations: (i) Limited work has been done to validate the use of thermal imaging across different skin tones [14, 15]. (ii) There has been no work comparing the effectiveness of thermal and optical imaging. (iii) Existing work trained on images acquired under strict data collection protocols where lighting, camera distance, and camera angle have been fixed. However, this is not practical in real-world practice.

The objective of our study is to address existing limitations by investigating the feasibility of thermography for detecting temperature differences in individuals with dark skin tones across various patient positions, lighting conditions, camera distance, and camera resolution. We collected optical and thermal images from 35 healthy adults (predominantly darker skin tones) and induced temperature differences on the lower back using two different protocols. We benchmark three CNN models using our dataset and evaluate the efficacy of detecting these temperature changes across diverse skin tones. The key contributions of our research are as follows: (i) Compilation of a diverse skin tone dataset (mostly focused on darker skin tones) comprising optical and thermal images from 35 patients; (ii) Comparison of pre-trained CNN model’s capability to detect temperature changes using either the optical or thermal images; (iii) Analysis of how the imaging protocol can impact the model performance in a controlled clinical simulation environment; and (iv) Assess the visual explanation of the CNN-based models to automatically identifying the region of interest without explicit bounding box labels.

## Method

### Data Collection

We collected optical and thermal images from 35 healthy adults in a controlled simulation environment. The study procedure was reviewed and approved by the Emory University Institutional Review Board, eIRB number 00005999. 30 participants had darker skin tones (Monk Skin Tone Scale level 6 or greater) and 5 had lighter skin tones (Monk Skin Tone Scale level 5 or lower). Using a digital colorimeter, we categorized the skin tone of subjects using the Eumelanin Human Skin Colour Scale [16] and had Eumelanin Intermediate Low (InterLow), Eumelanin Intermediate (Inter), Eumelanin Intermediate Mid (InterMid), and Eumelanin Intermediate High (InterHigh) categories as shown in Figure 1. The lower back of each participant was marked to conduct both a cooling and cupping protocol separately. The cooling protocol involved a cooled stone cylinder placed on the right lower back. The cupping protocol was performed using a suction device to induce erythema (i.e., abnormal redness of the skin).

**Figure 1:**
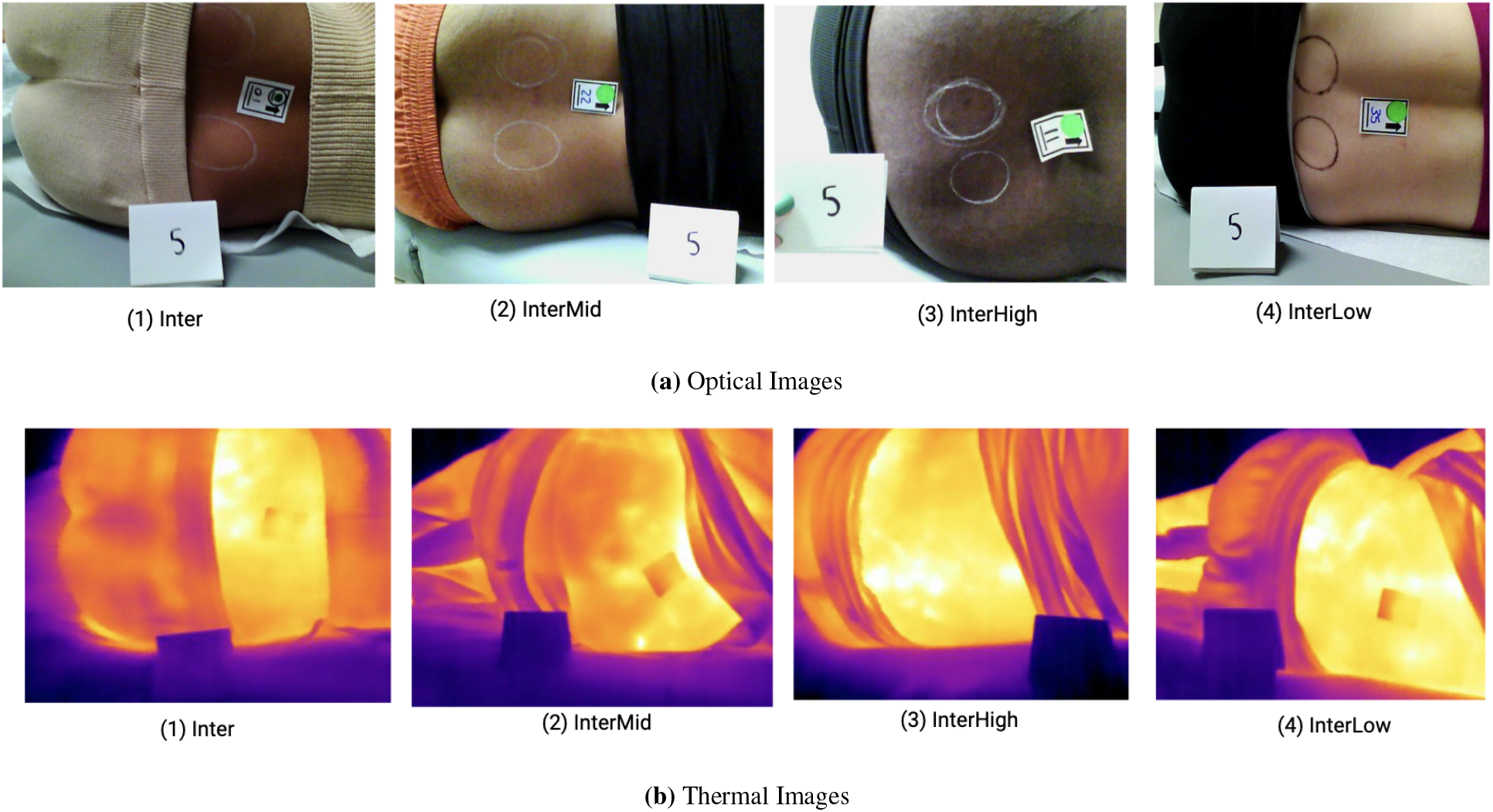
Example of control images from our dataset with the 4 different skin tones.

Images were taken with 2 thermal imaging cameras, the FLIR E8-XT (320×240 resolution) and the FLIR Pro One (160×120 resolution). For the cooling protocol, the imaging collection protocol was varied to capture (i) two lighting settings (ambient and ring light), (ii) two distances (35 and 50 cm), and (iii) three different postures (forward placement of top knee, knees stacked, backward placement of top knee). For each image acquisition setting (lighting, distance, posture) and camera, we captured a control and cool image pair (i.e., one before the cooling protocol and one after applying the stone cylinder). This yielded a total of 1680 images for the cooling dataset. For the cupping protocol, we used only the FLIR E8-XT at a single acquisition setting (50 cm and knees stacked). We collected a control image and 8 images within the first 7 minutes after the cupping device was removed (9 images per patient). Unfortunately, we were unable to acquire the erythema index for one of the images due to a laptop error, so there were a total of 314 images in the erythema dataset.

### CNN

CNNs have achieved remarkable performance in various medical imaging applications including dermatological assessments [17]. [11] proposed a CNN approach for classifying infrared thermal images, using 246 images from 82 patients with stringent imaging protocols. [12] used the MobileNetV2 Object Detection Model [18] to identify the PI location using thermal images from 10 subjects and 18 images using a tripod, specific distance, and dietary restrictions. Faster RCNN was also used to categorize the PI stage for 849 optical images [13]. Though these CNN models showed remarkable results, model generalizability may not be assured due to the small dataset sizes.

We used MobileNetV2, ResNet50, and InceptionNetV3 for our classification tasks. MobileNetV2 typically consists of an initial fully convolutional layer with 32 filters, followed by 19 residual bottleneck layers, and concludes with a 1×1 convolution, global average pooling, and a final fully connected layer [19]. The model’s efficiency and effectiveness make it a promising candidate for real-time PI detection, particularly in scenarios where computational resources may be limited, such as in portable devices used for bedside assessments [20].

ResNet50 is a CNN architecture that introduced the concept of residual learning to address the vanishing gradient problem in very deep networks [21]. It consists of 50 layers, including skip connections that allow the network to learn residual functions with reference to the layer inputs [22]. ResNet50 achieved state-of-the-art performance on the ImageNet dataset upon its introduction and has since become a popular backbone for various computer vision tasks, including image classification, object detection, and segmentation [23].

InceptionNetV3, also known as GoogLeNet, is another influential deep learning architecture designed for efficient image classification [24]. It employs “inception modules” that use multiple filter sizes and pooling operations in parallel, allowing the network to capture features at different scales simultaneously [25]. InceptionNetV3 improved upon its predecessors by incorporating factorized convolutions and auxiliary classifiers, which help with gradient flow during training [26]. This architecture has been widely used in transfer learning applications due to its strong performance and relatively low computational cost.

### Task Classification

We evaluate the efficacy of 3 image types for detecting temperature: an optical image, a black and white (B&W) thermal image, and a thermal color image. Thermal color images are the original temperatures captured directly by the thermal camera whereas the B&W thermal images are grayscale versions (0-255) derived from the color images. We assess the 3 image types on 2 tasks, cooling and erythema. Both of these are binary classification tasks. For the cooling task, we trained the model to identify whether the image was cool (+) or control (-). Similarly, for the erythema task, we classified the images as either positive if the measured erythema index is 6 c.u. above baseline (+), as measured with the colorimeter, or control (-). We randomly split the images into 80% training and 20% testing and ensured the image splits were the same across the 3 image types for fair comparison. For instance, if image 1 is in the training data of the optical dataset, the corresponding image 1 from both the thermal color dataset and the thermal B&W dataset will be included in their respective training data sets. After data splitting, we perform 5-fold cross-validation using the train data, with 4 folds for training and 1 fold for validation as shown in Figure 2. We evaluated performance on the test set using accuracy, sensitivity, specificity, F1, and area under the receiver operating characteristic curve (AUC).

**Figure 2:**
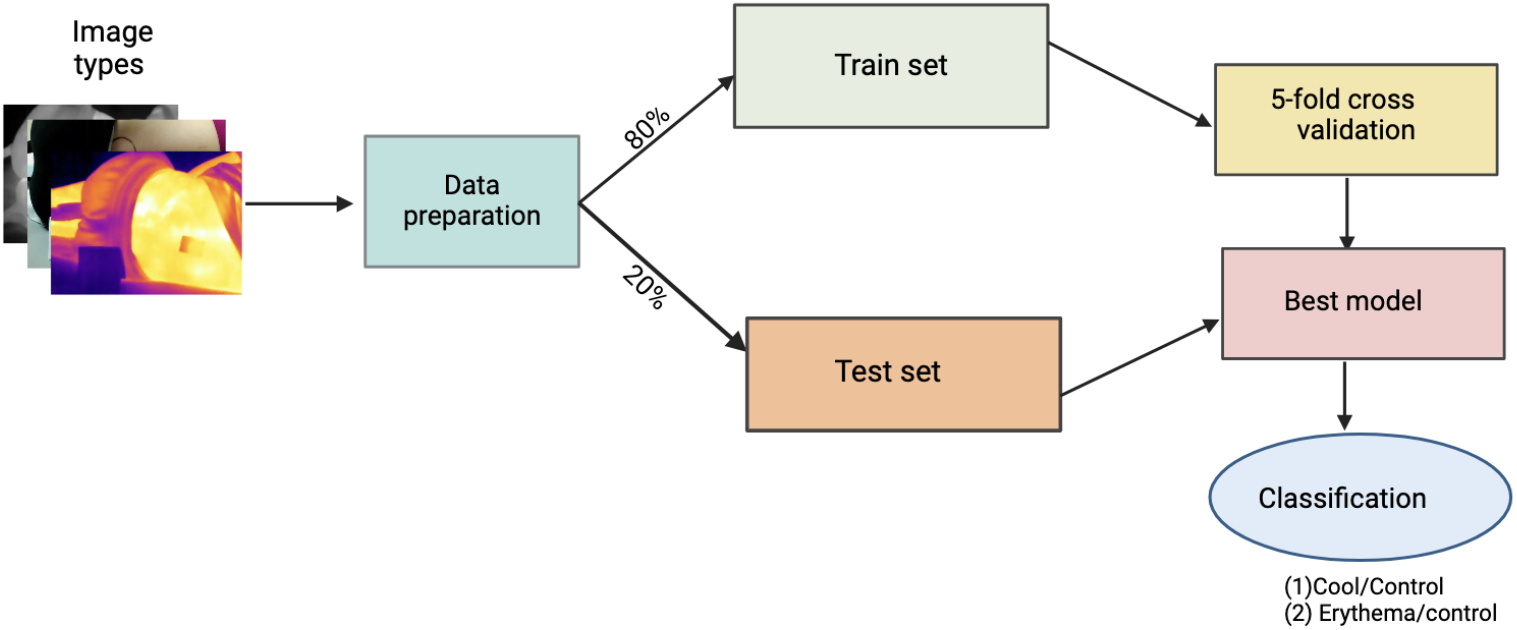
Our general approach to evaluating optical and thermal image classification using the CNN models.

### CNN Hyperparameter Tuning

All images for MobileNetV2 and ResNet50 were resized to 224×224, while those for InceptionNetV3 were resized to the default size of 299×299. We use 5-fold cross-validation to fine-tune all three CNNs for each image type and task. All 3 CNNs were pre-trained on ImageNet. We employed data augmentation to supplement our training set by incorporating either a horizontal flip or a 20-degree rotation. We fine-tuned each CNN with a batch size of 32, 50 epochs, a fixed learning rate, the Adam optimizer, and early stopping criteria. The model that achieved the highest validation accuracy over 5-fold cross-validation was selected and used to classify the images in the test set.

## Results

### Overall Predictive Performance

Table 1 summarizes the performance of MobileNetV2, InceptionNetV3, and Resnet50 on the 3 image types for the 2 tasks. For both tasks, thermal imaging (B&W and color) consistently outperforms their optical counterparts across many of the 5 metrics.

**Table 1:**
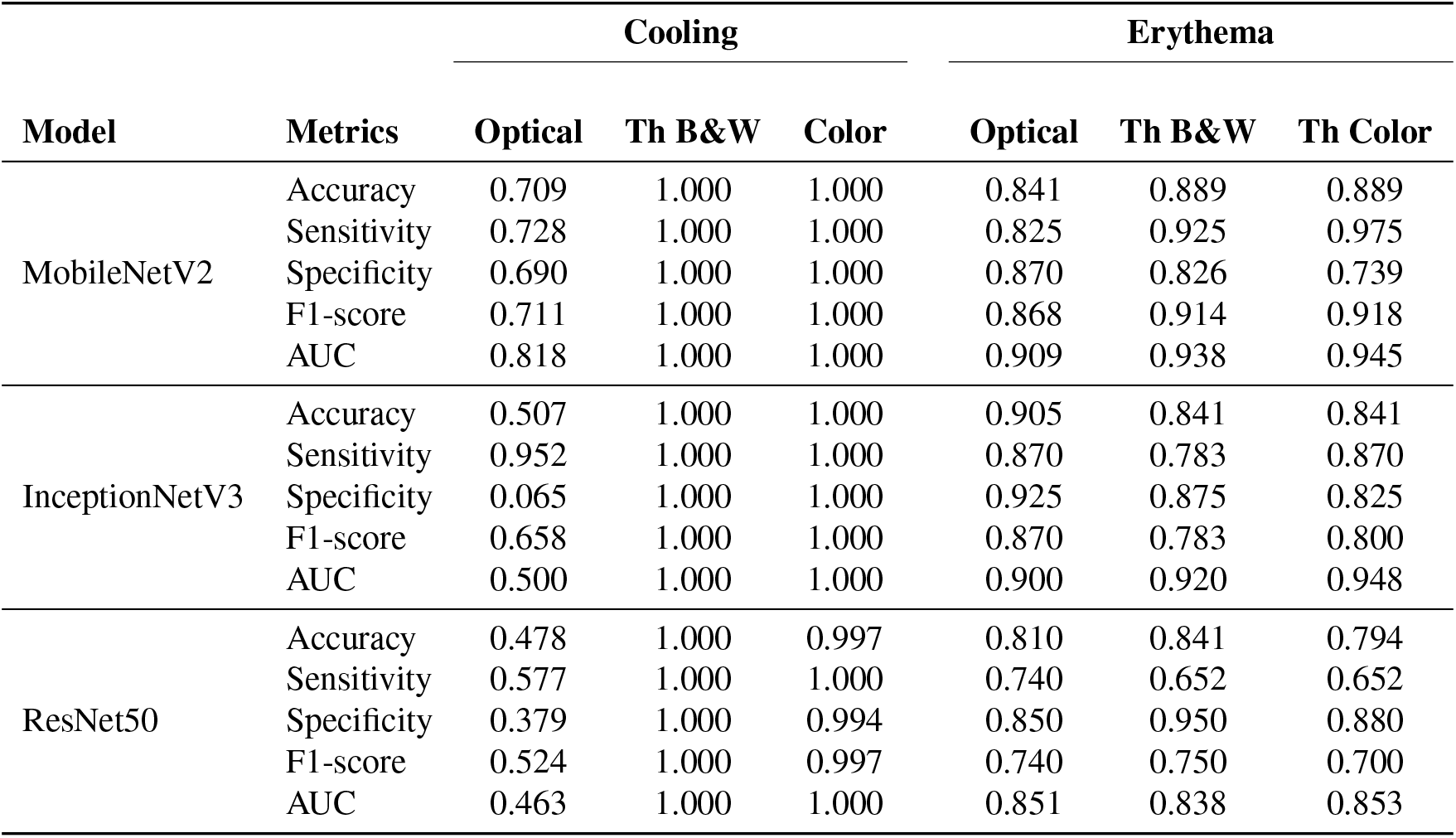
Comparison of the predictive performance results of the 3 CNN architectures for the 2 binary classification tasks, cooling and erythema, and the 3 image types (Th denotes thermal).

For the cooling task (an easier and more obvious temperature difference task), both thermal image types across all 3 achieve perfect performance with an AUC and F1 of 1, as opposed to 0.818 and 0.711 for MobileNetV2, 0.500 and 0.658 for InceptionNetV3, and 0.463 and 0.524 for ResNet50, respectively for the optical images. Notably, the larger models (ResNet50 and InceptionNetV3) did not yield better performance on the optical images suggesting some possibility of overfitting with the larger parameter spaces to the simpler models.

For the erythema task, the optical image achieved better performance than the cooling task with an F1 and AUC of 0.868 and 0.909 for MobileV2Net, 0.870 and 0.900 for InceptionNetV3, and 0.740 and 0.851 for ResNet50, respectively. This may be because erythema is accompanied by more visual color change than temperature change. For The thermal-based MobileV2Net generally achieves the best performance across the CNN models with F1 and AUC scores of at least 0.914 and 0.945 respectively. Notably, InceptionNetV3 achieves the highest AUC on the Erythema task of at least 0.948, but the F1 score is lower for both thermal imaging types than optical, as both sensitivity and specificity tend to be lower. For ResNet50, we observe that specificity tends to be higher for thermal, and thus F1 and AUC have slightly higher values for at least one of the thermal imaging types as opposed to the optical version. These results across both tasks suggest thermography is more reliable than optical imaging for detecting temperature changes may be more appropriate for diverse skin tones.

For the remainder of the results, we focus on the MobileNetV2 model as it consistently achieves the best performance across the 3 image types and both tasks. Figure 3 summarizes the confusion matrices for the cooling and erythema tasks for our fine-tuned MobileNetV2 across the different imaging modalities. These visual representations allow us to assess the accuracy and potential misclassifications in our models. With the cooling dataset, both thermal color and thermal B&W imaging demonstrate perfect classification, with 343 correct predictions out of 343 total cases (174 true negatives and 169 true positives) as shown in Figure 3a.

**Figure 3:**
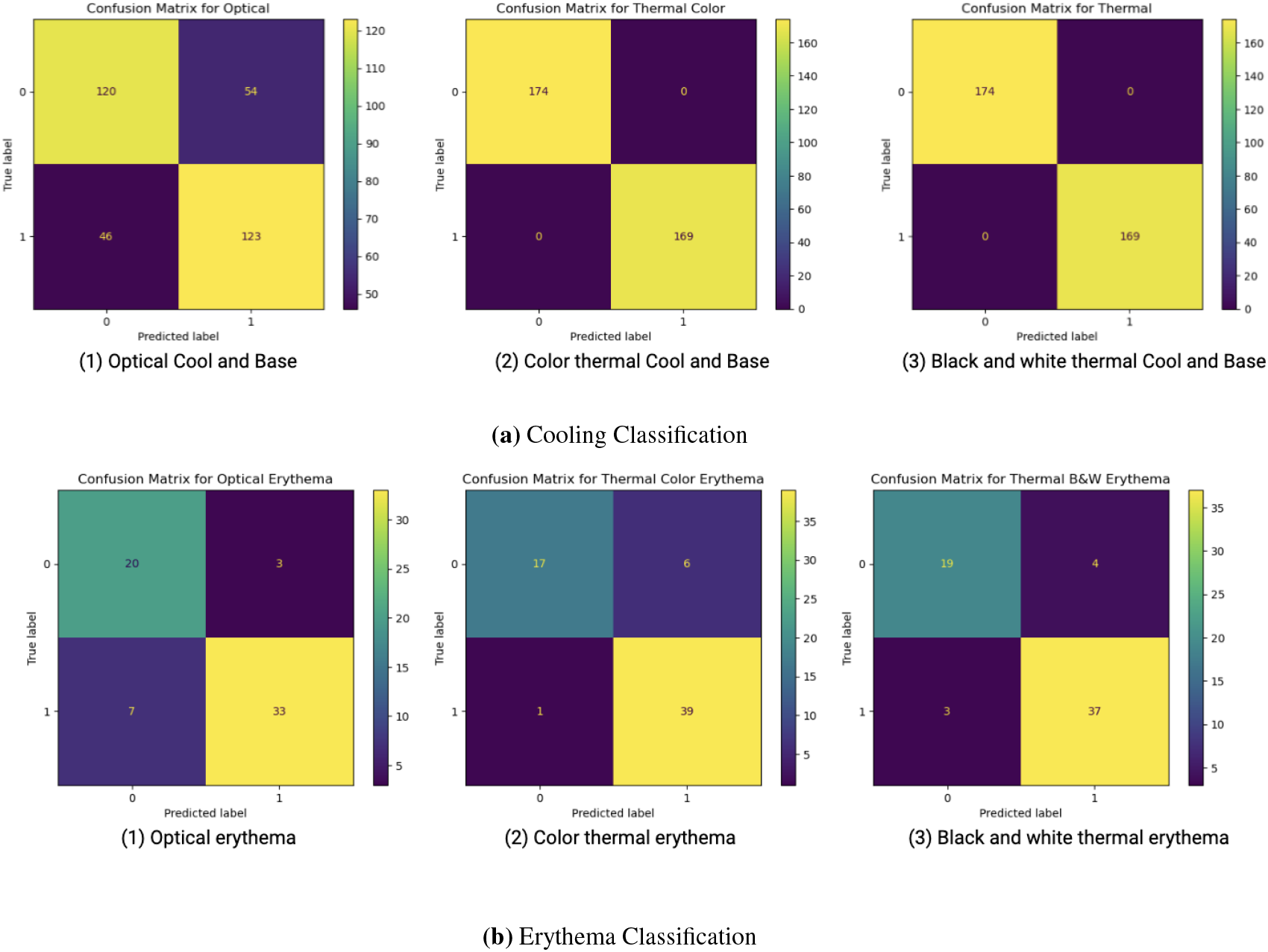
Confusion matrix from MobileNetV2 of the three image modalities on the 2 tasks.

The erythema dataset is also examined, comparing MobileNetV2 predictions against true labels for the presence or absence of skin redness. Among these, the thermal color imaging shows the highest overall accuracy, with 56 correct predictions out of 63 total cases (17 true negatives and 39 true positives) as shown in Figure 3b.

### Impact of Image Protocol

Table 1 suggests that thermography-based models are not sensitive to the image collection protocol, as the performance is stellar even with different lighting, camera distances, and patient poses. For the optical images, Figure 4 summarizes the errors associated with each of the 12 image collection protocols across the two cameras. These 12 protocols represent distinct combinations (hereby referred to as combo), each varying by knee position (Forward, Stacked, or Back), lighting condition (Room Light or Ring Light), and distance (35 cm or 50 cm). A higher number of images is an indication that a given protocol will perform poorly in detecting temperature changes, while a lower number of images indicates a potentially high performance associated with the collection protocol. The plot reveals that Combo 3 (Knee Forward, Ring Light, 35 cm) resulted in the highest number of misclassifications (12-13) while Combo 8 (Knees Stacked, Ring Light, 50 cm) had the lowest (about 5). The plot suggests that having the knees stacked together and taking the camera from a further distance (50 cm) generally yielded better performance. The ring light setting also was less ideal for when the top knee was forward or backward as it may create more shadows.

**Figure 4:**
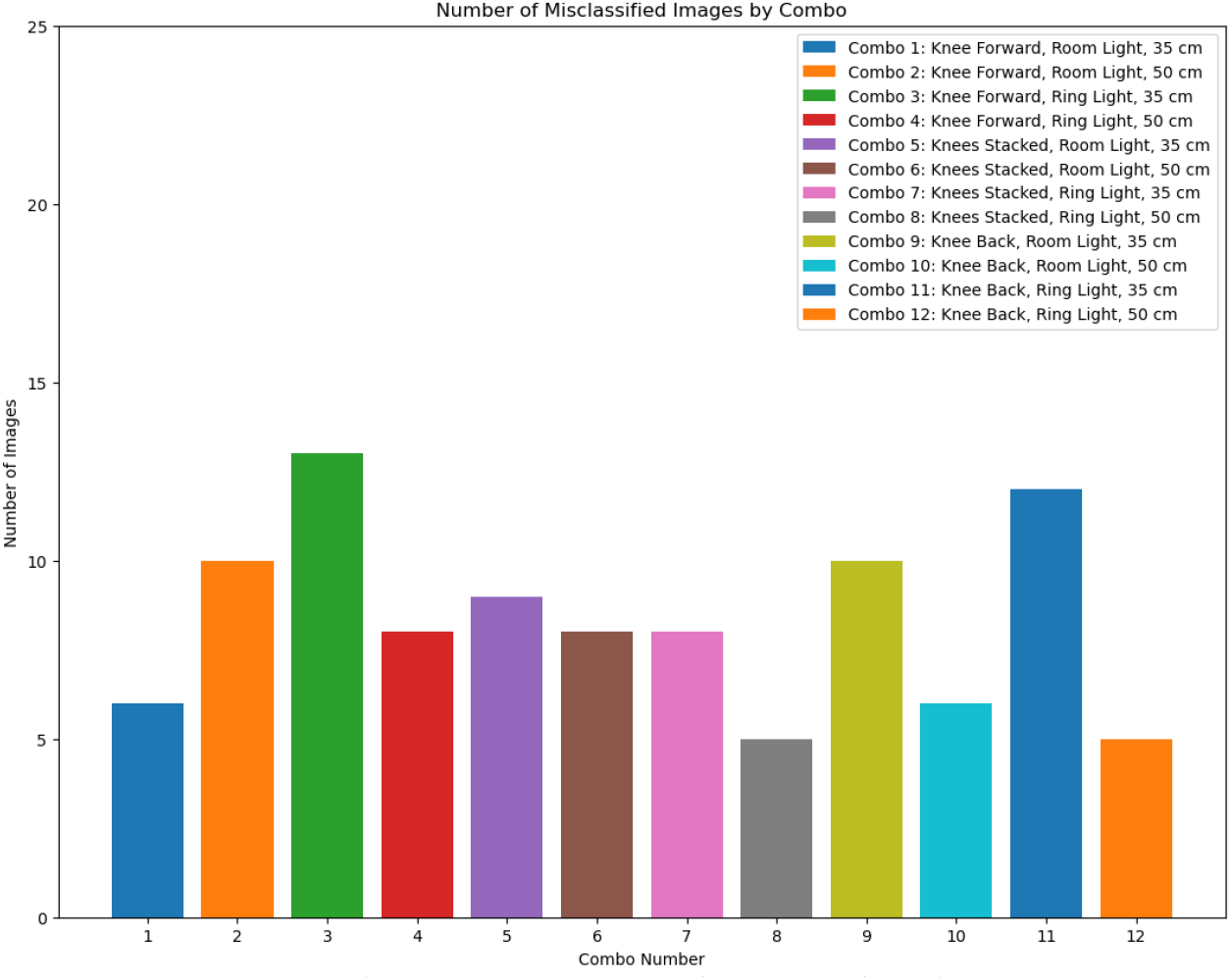
Bar plot of misclassified optical images on MobileNetV2 based on image collection protocol.

### Impact of Skin Tone for Erythema Detection

To better understand which skin tone categories are most susceptible to misclassification for the 3 image types, we further investigated the performance for each of the four skin tone categories. Table 2 summarizes the results of the test set for InterLow, Inter, InterMid, and InterHigh. As can be seen from the Table, many of the images were from the InterMid skin tone category. The results suggest that optical images perform slightly better for the Inter category (1 incorrect versus 2 for thermal imaging). However, thermal imaging is better for the InterMid category as there are 4 fewer mistakes in thermal imaging suggesting that for darker skin tones it is harder to detect redness in the skin.

**Table 2:** A comparison of the accuracy results using the MobileNetV2 architecture on the 3 image types, optical (Opt.), B&W thermal image, and color thermal image, for the erythema task by skin tone categories.

| Skin tone | Category | Optical | Th B&W | Th Color |
| --- | --- | --- | --- | --- |
| InterLow | Correct | 3 | 3 | 3 |
|  | Incorrect | 1 | 1 | 1 |
| Inter | Correct | 13 | 12 | 12 |
|  | Incorrect | 1 | 2 | 2 |
| InterMid | Correct | 32 | 36 | 36 |
|  | Incorrect | 8 | 4 | 4 |
| InterHigh | Correct | 5 | 5 | 5 |
|  | Incorrect | 0 | 0 | 0 |

### Interpretability of the model

Deep learning models are often considered ‘black boxes’ due to their complexity and lack of interpretability. To ensure the model’s transparency, techniques like Shapley Additive explanations (SHAP) [27] and Gradient-weighted Class Activation Mapping (Grad-CAM) [28] have been proposed. To better understand how MobileNetV2 was making its predictions and whether it necessitated providing a bounding box label for the region of interest, we used GRAD-CAM. Grad-CAM produces visual explanations for decisions made by CNN-based models. It uses the gradient of any target concept (e.g., ‘cat’ in a classifier) flowing into the final convolutional layer to produce a coarse localization map. This map highlights the important regions in the image for predicting the concept. We applied Grad-CAM to visualize the regions predicted by our model for all three image modalities (optical, color thermal, and B&W thermal) in both the cooling dataset and the Erythema dataset. From the GRAD-CAM visualization, it is evident that our fine-tuned MobileNetV2 is able to correctly detect the cool spot as shown in (1) and (3) in Figure 5. Moreover, the localized map is centered over the region of interest where the spot was placed for the thermal image (1) whereas for the optical cool, this seemed to be less consistent with stone placement location. Similarly, for the erythema task, MobileNetV2 is also able to correctly classify control images and provide reasonable explanations. For the control case, or image (2), the localization map was not focused on a specific region whereas for the erythema case, it correctly identifies the cupping region on the upper part of the back. This suggests that we may not need to provide the bounding region of interest, which was integral for many of the existing CNN-based PI models performing image segmentation of the thermal images.

**Figure 5:**
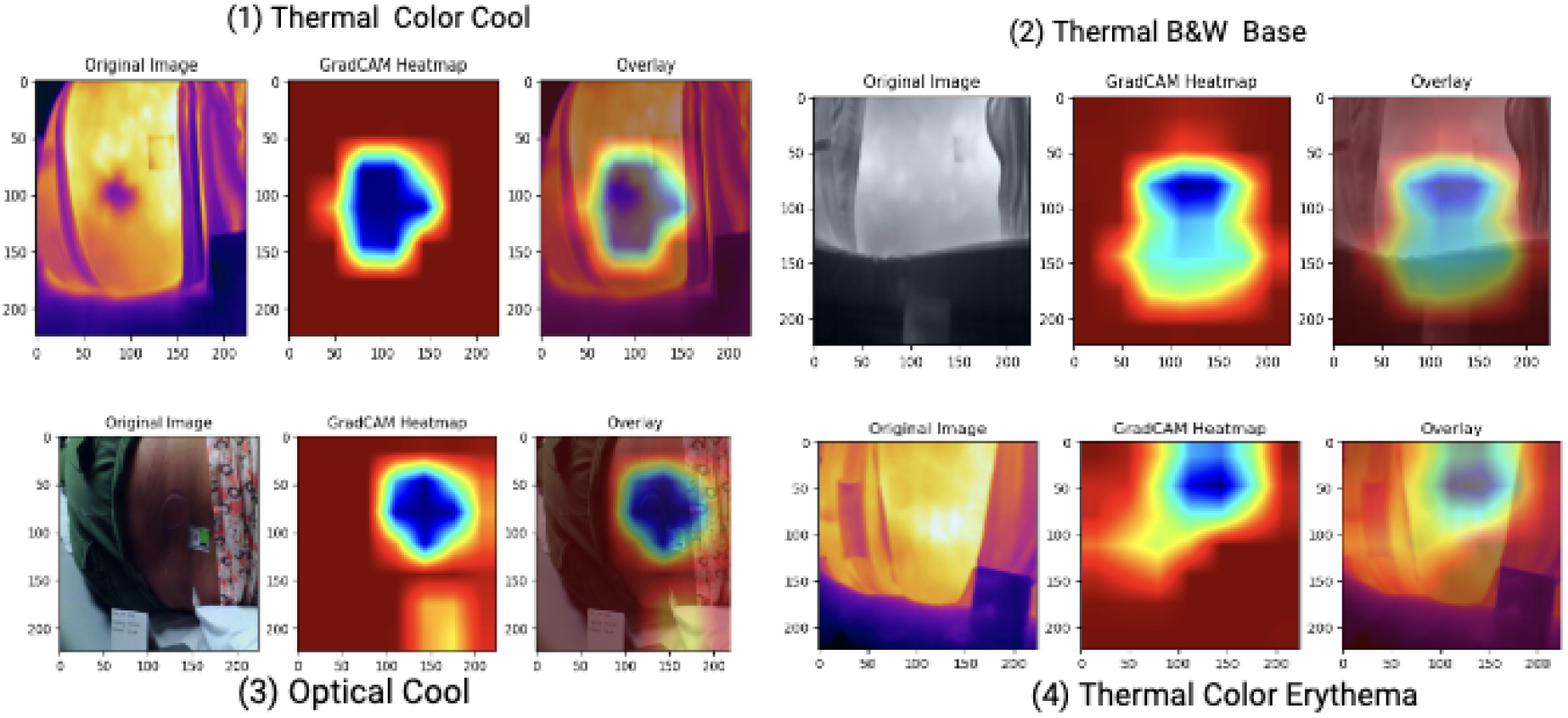
GRAD-CAM visualization of classifying cool, control (base), and erythema images using MobileNetV2 model.

### Impact of Augmentation on Model Performance

We examine the effect of data augmentation on MobileNetV2’s performance using the thermal color erythema images. The impact of augmentation was explored because it will be infeasible to follow a strict imaging protocol for PI assessment in a typical clinical setting. Two augmentation techniques, horizontal flip and rotation at 20^*°*^, were applied to both train and test images. We train two versions of the MobileNetV2, one with the augmented training images, and one without the augmented training images (i.e., the original collected training images only). We evaluate the performance on both the original test set without data augmentation and the test set with augmented data images.

As shown in Table 3, training with augmentation yielded the best AUC for both test sets (with and without data augmentation). However, training without augmentation had the best F1, accuracy, and specificity on the test without data augmentation. Notably, training without augmentation yielded substantial performance differences between the augmented and non-augmented test set with the F1 score dropping by almost 0.10 points and specificity falling from 0.826 to 0.261. In contrast, training with augmentation yielded somewhat similar scores on the two sets in terms of F1-score and AUC. These results imply that augmenting training images does not negatively impact model performance, and in fact, makes it more robust to a general setting. This may indicate that data augmentation can be used to develop an accurate PI prediction model that is capable of handling the varying nature of images that are captured in a typical clinical setting.

**Table 3:** Comparing the results of augmenting training and test images on the performance of thermal erythema image classification using MobileNetV2 where aug denotes augmentation.

| Metrics | Training with aug |  | Training without aug |  |
| --- | --- | --- | --- | --- |
|  | Test with aug | Test without aug | Test with aug | Test without aug |
| Accuracy | 0.889 | 0.889 | 0.730 | 0.905 |
| Sensitivity | 0.975 | 0.975 | 1.000 | 0.950 |
| Specificity | 0.739 | 0.739 | 0.261 | 0.826 |
| F1-score | 0.918 | 0.918 | 0.825 | 0.927 |
| AUC | 0.927 | 0.945 | 0.823 | 0.944 |

## Discussion

Our findings demonstrate the potential of thermography and CNNs to detect temperature changes in individuals with darker skin tones more reliably than optical images. Although the MobileNetV2 performance is slightly lower for detecting erythema in slightly darker skin (InterMid skin tone category), the results are still better than their optical counterparts. This suggests that thermography may be a viable solution for detecting PI even on dark skin tones – a significant challenge under current clinical practices. Our preliminary results also suggest that thermography does not require adherence to a strict imaging protocol as there was little performance difference between the patient pose, lighting setting, and camera distance. This may serve as a crucial finding as following a stringent imaging protocol may adversely impact nursing workflow and impede adoption.

Our preliminary findings provide promising implications for the adoption of thermography for PI detection even for diverse skin tones. However, we do note that there are several limitations to our current work. First, our patient sample is quite small (35 patients) with many from the InterMid skin tone category. As such, a larger sample size is necessary to draw more substantive conclusions. Second, the images were collected in a controlled and simulated setting and did not include real PI. Although there is some evidence that thermography is fairly robust to lighting, pose, and distance, this may change for real-time deployment as patients may not be capable of lying on their side or nurses may not provide high-quality images.

## Data Availability

The thermography data in the present study is available upon reasonable request to the authors. The optical images may contain patient-identifiable information and will require IRB approval for access.

## Acknowledgements

This work was supported in part by the National Center for Advancing Translational Sciences of the National Institutes of Health under Award number UL1TR002378. The content is solely the responsibility of the authors and does not necessarily represent the official views of the National Institutes of Health.

